# Cost-Effectiveness of a Risk-Based Pancreatic Cancer Early Detection Strategy in New-Onset Diabetes Patients Using a Blood-Based, Non-Invasive Test – An Economic Analysis

**DOI:** 10.1101/2025.11.12.25340092

**Authors:** Adrian Vilalta, Ananya Das, Michael B. Wallace

**Affiliations:** ClearNote Health, San Diego, CA; Outcomes Metrics LLC, San Diego, CA; Arizona Centers for Digestive Health, Phoenix, AZ; Mayo Clinic, Jacksonville, FL

## Abstract

This study aims to assess the clinical and economic benefits of early detection of pancreatic cancer (PC) in patients with new-onset type 3c diabetes (NOD) using a blood-based, cell-free DNA epigenomic test (Avantect; ClearNote Health, CA). Multiple studies indicate that NOD increases the risk of PC by 6-8 times within the first three years post-diagnosis.

A Markov model was created to compare two primary strategies: no testing and testing higher-risk NOD patients using a blood-based cell-free DNA epigenomic test. Additionally, imaging surveillance strategies were also evaluated. Using criteria from Sharma *et al*. (*Gastroenterology*, 2018), approximately 20% of the NOD patients were considered to be at higher risk for PC. The risk for developing PC, survival, and cost data were obtained from the US Surveillance, Epidemiology, and End Results (SEER) and Medicare databases.

The cell-free DNA epigenomic test has a sensitivity of 68% and a specificity of 97%. It proves robustly cost-effective in NOD high-risk patients, with an Incremental Cost-Effectiveness Ratio (ICER) of $56,564 at a Willingness to Pay (WTP) of $100,000

The model predicts that in a cohort of 10,000 patients with NOD, 1% would be diagnosed with PC in the no-testing strategy, with only 7.1% of cases eligible for surgery. In contrast, 71 PC cases are predicted to be detected with cfDNA testing. These cases would be more likely to be detected at an earlier, more treatable stage, with 32.4% eligible for surgical resection.

Testing higher-risk patients with NOD for pancreatic cancer using a blood-based cell-free DNA epigenomic test is predicted to be cost-effective compared to the standard of care (no testing). Initiating this test within three years of a diabetes diagnosis is likely to increase the detection of treatable pancreatic cancer cases, potentially improving patient survival outcomes.

**KEY POINTS:** *Question:* What are the economic benefits of testing for pancreatic cancer (PC) in patients with New-Onset Diabetes using a cfDNA epigenomic test?

*Findings:* Markov model predicts that of 10,000 hypothetical patients with NOD, 1% would be diagnosed with PC in the no-surveillance strategy, with only 7.1% of cases treatable with surgery. Meanwhile, 71 PC cases are predicted to be detected with cfDNA testing. The latter cases are predicted to be detected at an earlier, more treatable stage, with 32.4% eligible for surgical resection.

*Meaning:* cfDNA testing of risk-stratified New-Onset Diabetes patients within 3y of diabetes diagnosis is cost-effective.

## INTRODUCTION

Pancreatic cancer (PC) is a leading cause of cancer mortality around the globe. PC is the third most common cause of cancer-related deaths in the US, and it is predicted to be the second-leading cause by 2030.^1^ In the US, the 5-year survival expectancy after diagnosis is only 13.3%.^1^ However, the stage at which cancer is detected determines the treatment options and subsequent survival expectancy. Currently, over 50% of PC cases are diagnosed as “distant,” with a 5-year survival rate of roughly 3% for such advanced disease cases.^1^ On the other hand, detecting the disease at stage IA improves the expected 5-year survival rate to approximately 80%.^2^ Therefore, early detection is vital to reducing the disease burden of pancreatic cancer.^3^

Currently, the initial evaluation of patients for PC relies primarily on imaging-based testing modalities.^4^ Serum biomarkers, such as CA19-9 and the carcinoembryonic antigen (CEA), are often used with imaging to evaluate symptomatic patients.^4^ However, imaging approaches are not suitable for testing a large number of individuals, and CA19-9 and CEA are neither sensitive nor specific enough to be used for the early detection of PC.^5^

According to US professional guidelines, testing individuals for PC is currently only recommended for individuals at increased risk, as defined by family or genetic history or known cystic neoplasms of the pancreas.^6-8^ In such individuals, annual MRI or endoscopic ultrasound imaging (EUS) is standard and has been demonstrated to be cost-effective.^9^ However, there is a need for studies in the broader population of high-risk individuals, such as those with New Onset Diabetes type 3.

Sensitive, accurate, non-invasive testing could address the need for PC early detection when imaging modalities cannot be used routinely. Analysis of cell-free DNA (cfDNA) found in blood has proven to be a powerful platform for developing non-invasive testing. Data first published by Guler *et al*. show differences in the distribution of the epigenomic marker 5-hydroxymethylcytosine (5hmC) in cfDNA samples from patients with PC compared to healthy individuals across various genome regions.^10^ The authors used cfDNA isolated from patients’ peripheral blood to determine 5hmC-DNA profiles using a proprietary 5hmC enrichment workflow and Next-Generation Sequencing (NGS). These epigenomic profiles were used to train a machine-learning algorithm that can detect pancreatic cancer at an early stage. These findings have been validated, demonstrating that the test can detect PC at an early stage with high sensitivity and specificity.^11,12^ This technology is the basis for the commercially available Avantect Pancreatic Cancer Test (cfDNA test).

Type 2 diabetes (T2D), and especially new-onset type 3c (NOD), is a significant risk factor for pancreatic cancer. Type 3c diabetes, also called pancreatogenic diabetes, occurs when damage to the pancreas reduces insulin or glucagon production.^13^ However, the differential diagnosis of type 3c diabetes can be challenging, and it is therefore often misdiagnosed as type 2 diabetes.^13^ Multiple retrospective studies have demonstrated an elevated risk of PC within 2 to 3 years of NOD diagnosis, an association confirmed across racial and ethnic groups.^14^ Nearly 25% of patients with PC are diagnosed with diabetes six months to 36 months before the diagnosis of PC.^15^ Individuals 50 years of age and older with glycemically defined NOD have a 6-8-fold higher risk of a pancreatic cancer diagnosis within three years of meeting the criteria for NOD.^16^ The 3-year incidence of PC in this group is approximately 1%.^16^ Analysis of an extensive real-world database of administrative claims has further confirmed the association of NOD with a subsequent PC diagnosis.^17^ Importantly, recently published prospective data from a large NOD cohort further confirm these findings.^18^ Therefore, testing for cancer at the time of diabetes diagnosis may lead to the identification of PC at an earlier stage, when treatment may significantly increase the chances of survival.^16,19^ However, despite the risk of PC conferred by NOD, current management of a new diabetes diagnosis does not include testing for PC.

Economic models evaluating the potential cost-effectiveness of risk-based surveillance of NOD patients using imaging modalities have been published.^20,21^ Our study aims to extend the understanding of the economics of PC testing by developing a cost-effectiveness model for PC detection using a blood-based, non-invasive cfDNA test (Avantect Pancreatic Cancer Test).

## METHODS

We developed a Markov model to simulate the risk of developing pancreatic cancer in individuals newly diagnosed with diabetes (NOD), adhering to the guidelines of the Panel on Cost-Effectiveness in Health and Medicine for conducting and reporting a reference case analysis.^22,23^

For the initial analysis, we evaluated competing strategies in 50-year-old cancer-free NOD patients from the viewpoint of a third-party payer, considering a lifetime time horizon.

### Study Definitions

NOD was identified in patients who had two consecutive or simultaneous hyperglycemia indicators (fasting blood sugar ≥ 126 mg/dl, random blood sugar ≥ 200 mg/dl, and hemoglobin A1c ≥ 6.5) without previous diabetic treatment and who had at least one normal glucose test in the prior 3-18 months. Surveillance candidacy in these patients was determined using an END-PAC score of ≥ 3.^19^ We did not consider patients with high-risk familial or genetic susceptibility for pancreatic cancer.

### Markov Model and Clinical Probabilities

We constructed a Markov model with six transitional states: NOD, low-risk lesion, high-risk lesion, pancreatic cancer, diabetes, and death, as depicted in **Figure 1**. This model was converted into a decision tree using TreeAge Pro software (TreeAge Software, Inc., Williamstown, Mass). We derived clinical probabilities from various sources, including the SEER database for pancreatic adenocarcinoma distribution and a systematic review of surveillance test results (EUS or MRI)(**Table 1**). Mortality data during pancreatic cancer treatment also came from SEER, while baseline complication probabilities were sourced from published studies.^9,24^

**Table 1:**
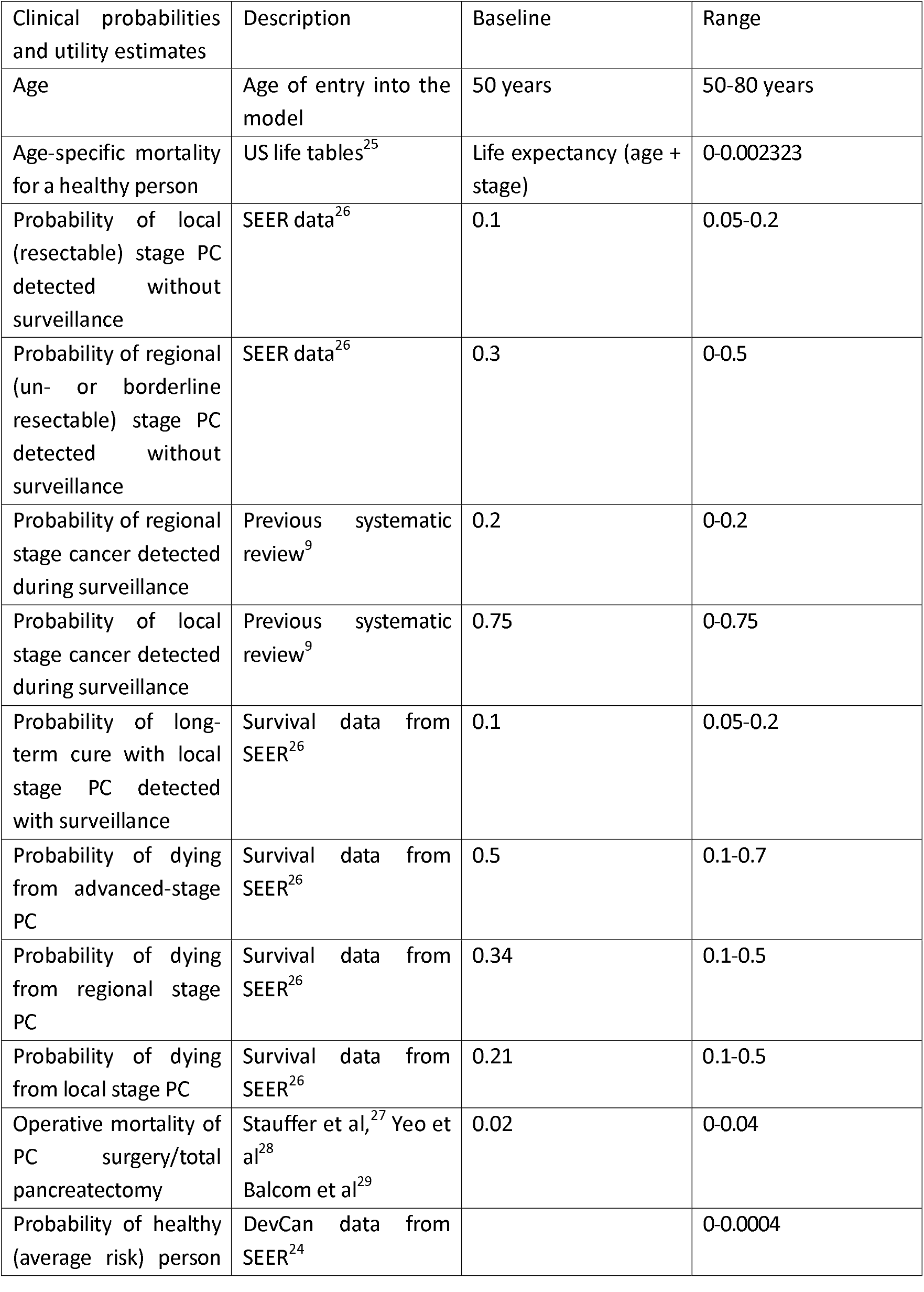
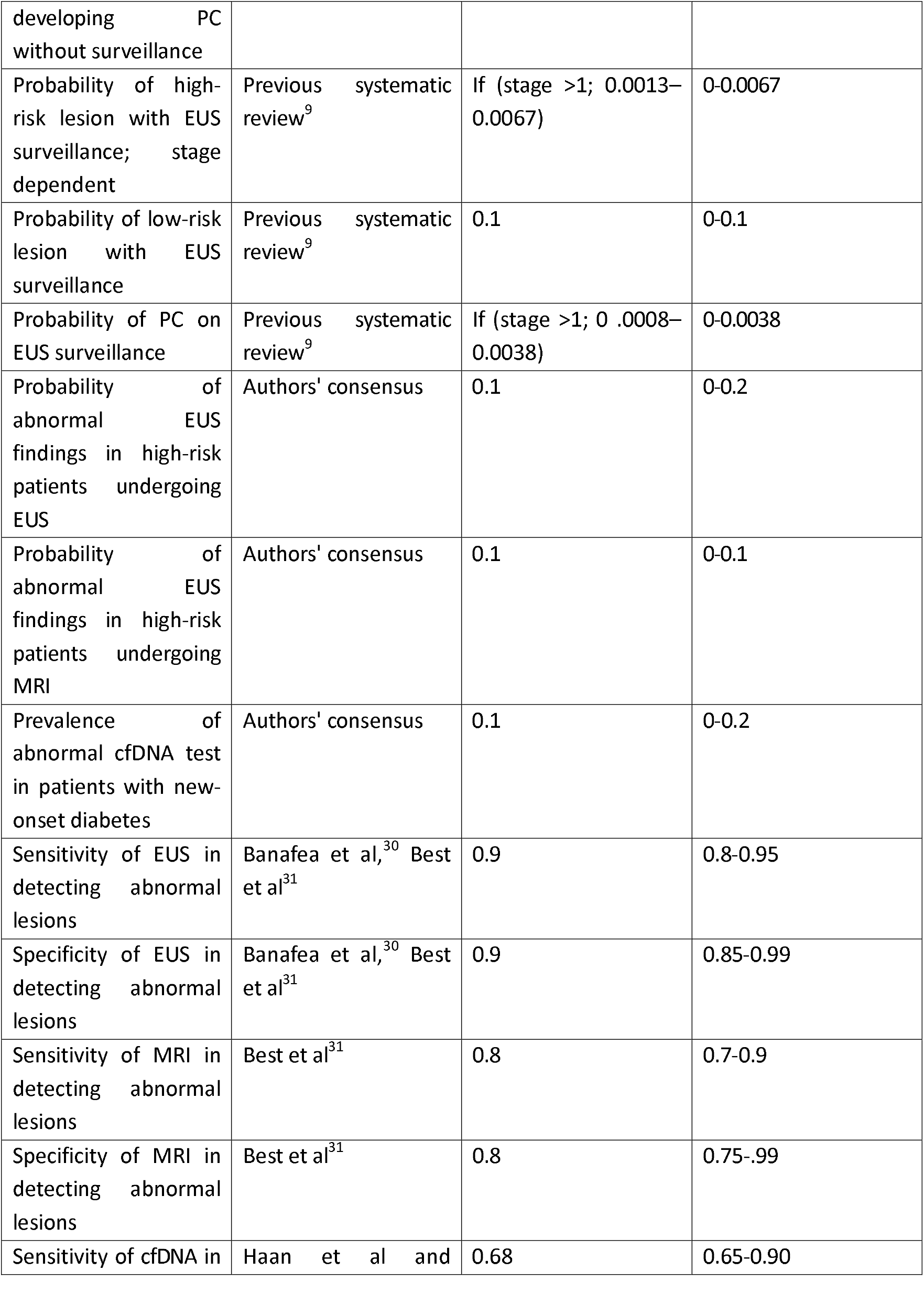
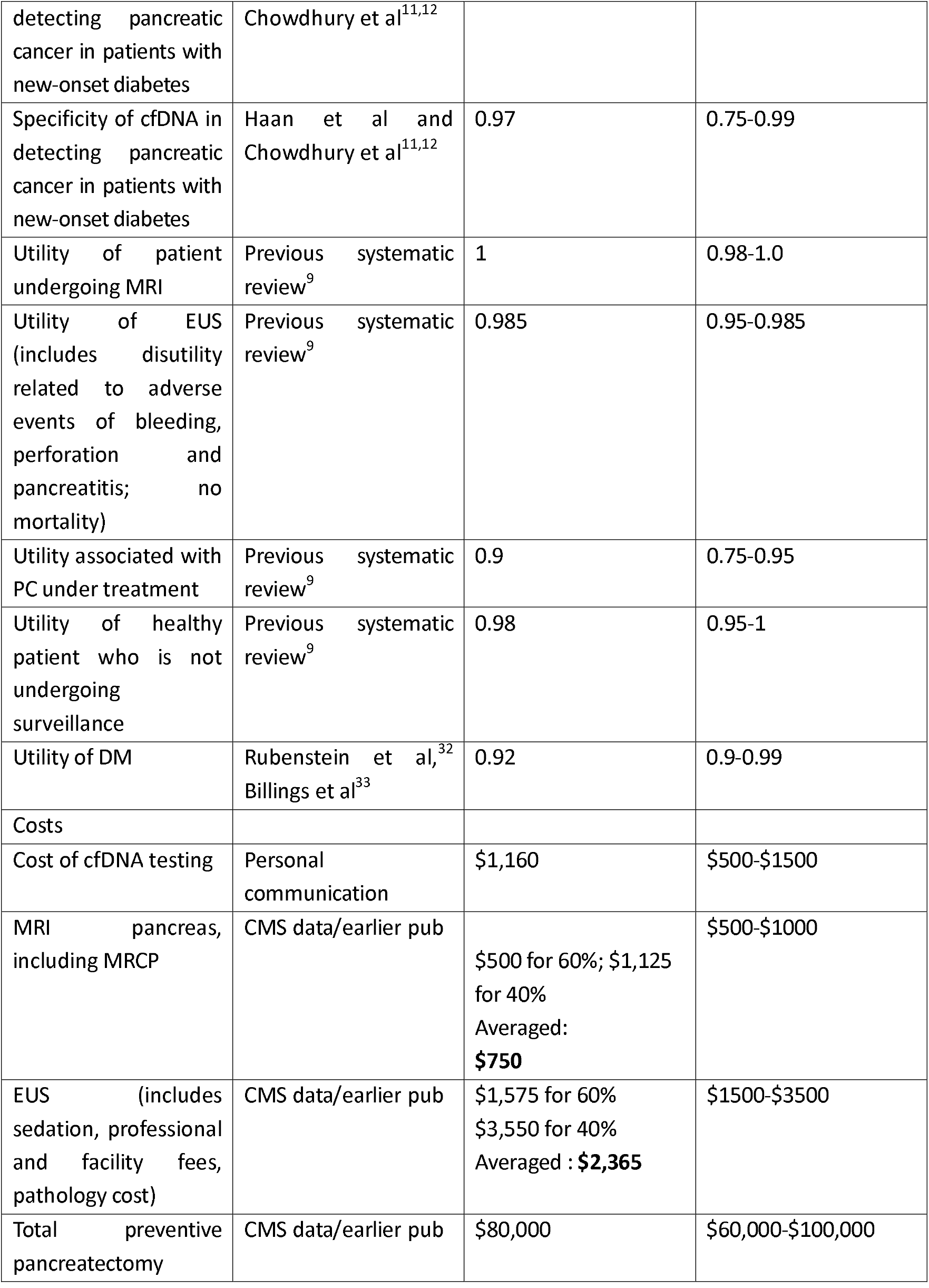
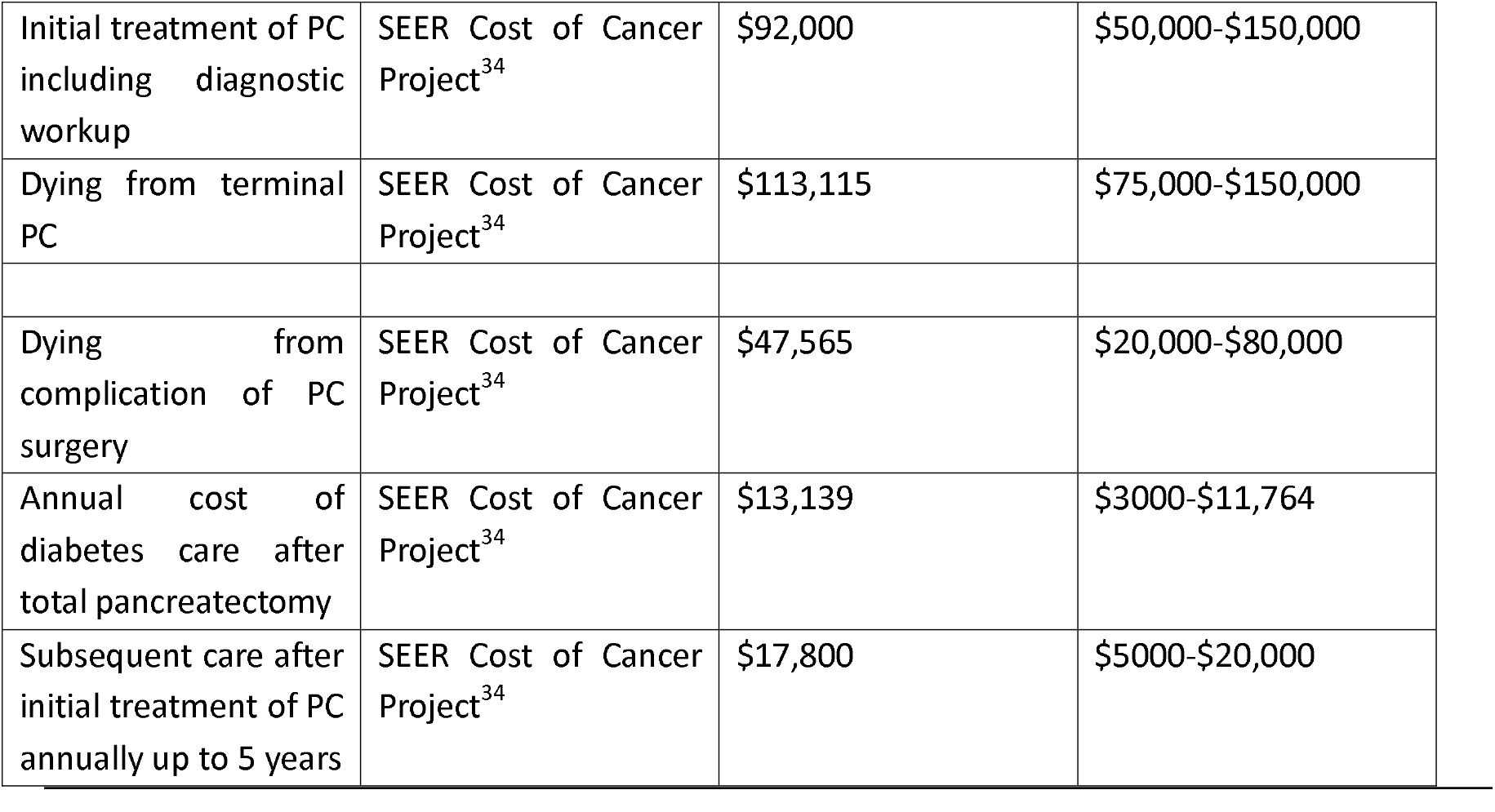
Selected clinical probabilities and cost estimates used in the decision analysis (modified from previous publication^9^)

**Figure 1.**
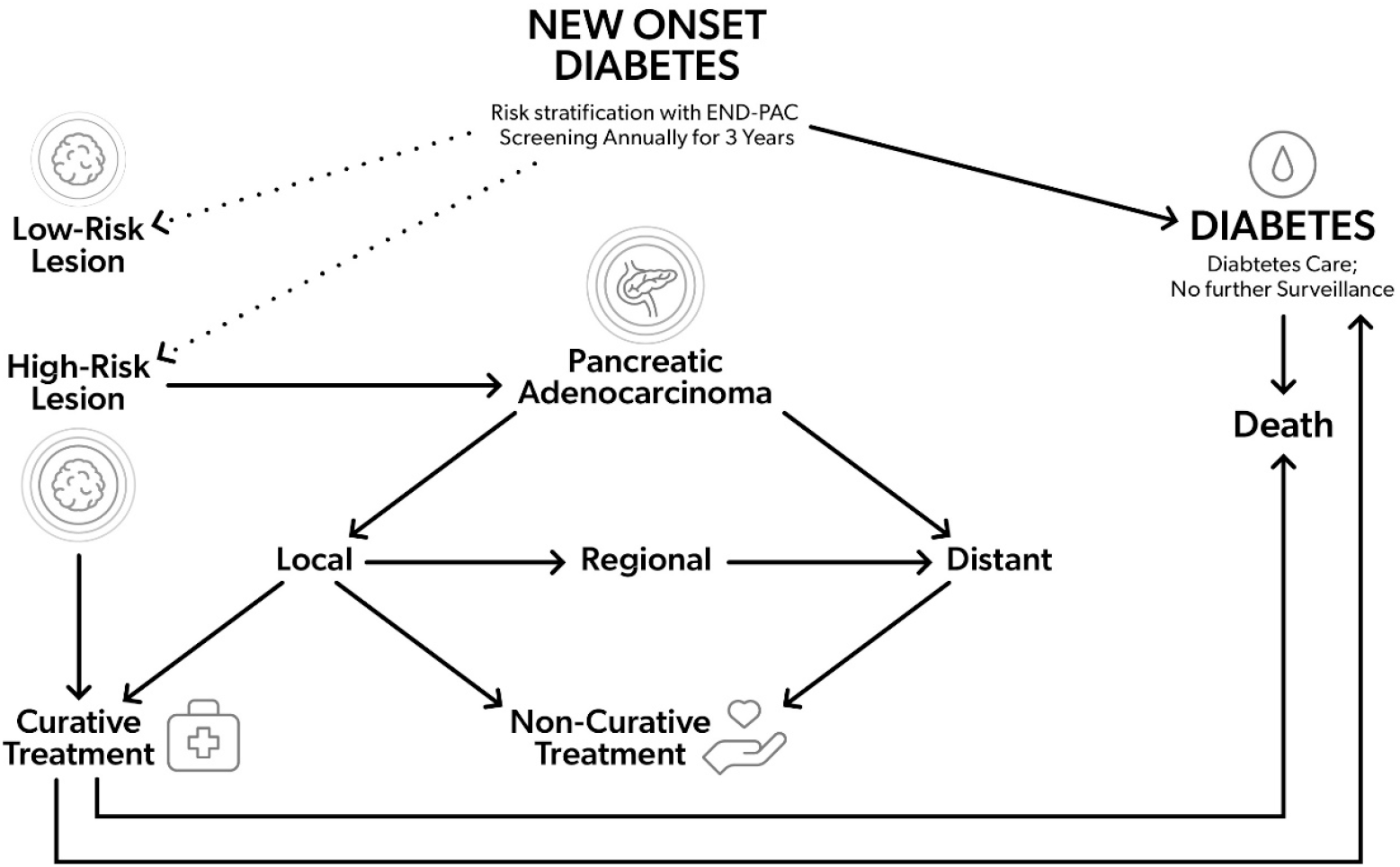
The figure shows a simplified model of different Markov states. Patients with new-onset diabetes remain in the diabetic state and undergo surveillance tests annually, stratified by the Mayo END-PAC risk score. A minority of these patients will develop either low-risk or high-risk lesions or transition into different stages of pancreatic cancer and undergo stage-specific curative (surgery) or non-curative treatment. If surgery is performed, the patient will transition through the perioperative state; if surgical resection is not feasible, they will be considered inoperable and continue to be in that state or may die in future cycles. Death is allowed from any state, but specific transitions are not allowed. A single arrowhead indicates a transition from one state to another in the direction of the arrowhead.

### Surveillance Strategies

We compared five surveillance strategies in the baseline analysis, applied after entry into the decision model:

- Strategy I, No Surveillance.
- Strategy II, MRI-based Surveillance with once-a-year MRI for 3 years.
- Strategy III, cfDNA-based Surveillance with once-year testing for 3 years.
- Strategy IV,EUS-based Surveillance with once-a-year EUS for 3 years.
- Strategy V, all patients with NOD had cfDNA testing at entry into the model; if the cfDNA testing was negative, no further testing was undertaken; if the test was positive and the patient had an END-PAC score of < 3, an initial EUS followed by cfDNA testing was done for 3 years; if the cfDNA test was positive and the patient had an END-PAC score of ≥3, annual EUS-based surveillance was continued for 3 years.

Surveillance commenced upon diagnosis of diabetes in individuals aged 50 or above and continued for three years. In strategies II to IV, patients with an END-PAC score of less than 3 were not tested. Smoking status did not affect the initiation of surveillance. CT scans were not considered for surveillance in accordance with CAPS guidelines. In strategies II through V, if imaging showed a low-risk lesion, EUS-guided fine-needle aspiration (EUS-FNA) was performed. If the EUS-FNA result is negative, EUS would be repeated in 6 months. If a high-risk lesion or cancer were found on surveillance and were confirmed by EUS-FNA, surgery would be performed. All patients enrolled in the surveillance program were considered surgical candidates.

### Cost Estimates

Cost data were primarily sourced from the SEER database and the Centers for Medicare & Medicaid Services (CMS), with adjustments made for geographic cost variations. (Table 1) We considered direct costs only, adjusted for inflation and discounted at an annual rate. An inflation rate of 3% and a discount rate of 3% were applied annually.

CMS refers to the Centers for Medicare & Medicaid Services; MRCP stands for magnetic resonance cholangiopancreatography.

### Outcomes Estimates

Outcomes were measured in quality-adjusted life years (QALYs), with incremental cost-effectiveness ratios (ICERs) calculated considering expected lifetime quality adjustments for the diabetic state and postoperative conditions. The postoperative state was calculated using utility values of short-term states (e.g., chronic abdominal pain, medication intake [pancreatic enzyme replacement], and diabetes).^9^ Mortality from surgery or cancer was accounted for by assigning a utility value of 0.

### Sensitivity and Threshold Analysis

We tested model robustness using sensitivity and threshold analyses to assess the impact of various clinical probabilities and cost factors. A probabilistic sensitivity analysis was conducted in a simulated cohort of 10,000 patients, enabling us to estimate risk reductions and determine necessary surveillance measures. Sensitivity analysis was performed with the performance characteristics of cfDNA testing and imaging tests (i.e., EUS, MRI, or a combination of both), costs, surveillance interval (i.e., every year vs every three years), and surveillance duration. Threshold analysis determined a cost limit under which EUS and MRI stopped being cost-effective. Similarly, we calculated an age limit at which surveillance costs exceeded QALYs gained.

### Willingness to Pay (WTP) and Net Health Benefit

We assessed cost-effectiveness across a Willingness-to-Pay (WTP) range of $100,000 to $200,000, using net health benefit calculations as an alternative to ICERs.

### Statistical Considerations and Other Assumptions

We incorporated half-cycle corrections and assumed diminished follow-up test positivity after an initial negative result. Sensitivity and specificity were considered for cfDNA testing and both EUS and MRI. For pancreatic cancer patients who had advanced disease, treatment costs were limited for 1 or 2 years, followed by death. Expertise was assumed for both procedural and interpretive tasks, and surgeries were restricted to high-volume centers, opting for total pancreatectomy to preempt further cancer development. Any discrepancies in the data were resolved through consensus among the authors.

## RESULTS

Baseline analysis of a cohort with new-onset diabetes with a Mayo END-PAC score of ≥ 3 showed that the cfDNA-based surveillance strategy is cost-effective compared to the no-test strategy at an ICER of $56,564, which is well *below the usual willingness to pay threshold (WTP, $100, 000-$200,000*). Our analysis showed that the cfDNA-based strategy yielded a higher QALE compared to the MRI-based strategy over the no-test strategy. In a direct comparison of the cfDNA-based strategy with MRI-based surveillance, although the MRI-based strategy was less costly, it yielded less QALE. EUS-based surveillance was prohibitively expensive, with an ICER of almost $ 122 million. CfDNA testing done at entry, without considering the Mayo END-PAC score, was absolutely dominated, as it was more expensive and yielded fewer QALYs compared to no surveillance. The results of the baseline cost-effectiveness analysis are shown in **Table 2**.

**Table 2:** Results of the baseline analysis.

| Strategy | Cost | Incremental Cost | Effectiveness (QALE) | Incremental Effectiveness (QALE) | ICER (\$/QALE) | NMB |
| --- | --- | --- | --- | --- | --- | --- |
| No Testing | \$169,861 | | 11.75006 | | | \$2,180,150 |
| MRI | \$170,374 | \$513 | 11.79719 | 0.04713 | 10,881.00 | \$2,189,065 |
| cfDNA | \$170,576 | \$202 | 11.80076 | 0.00357 | 56,564.00 | \$2,189,577 |
| EUS | \$219,964 | \$49,388 | 11.80117 | 0.00040 | 22,412,529.00 | \$2,140,269 |
| cfDNA-all NOD | \$228,226 | \$8,262 | 11.79503 | -0.00613 | -1,347,170.00 | \$2,130,781 |

### Sensitivity and Threshold Analyses

Sensitivity analyses confirm that although the model’s results are influenced by the costs and performance characteristics of the tests, the cfDNA-based strategy remains cost-effective within the range documented in the literature. A tornado diagram illustrates that this strategy withstands variations in most modeled parameters. (**Figure 2**) The age of entry into the model significantly impacts cost-effectiveness, with the cfDNA-based surveillance maintaining affordability across a broad age range (40 to 80 years). Notably, between the ages of 64 and 72, this strategy is unequivocally dominant—both less expensive and more effective. Interestingly, MRI-based surveillance only falls below the willingness-to-pay threshold between ages 69 to 71. (**Figure 3**)

**Figure 2.**
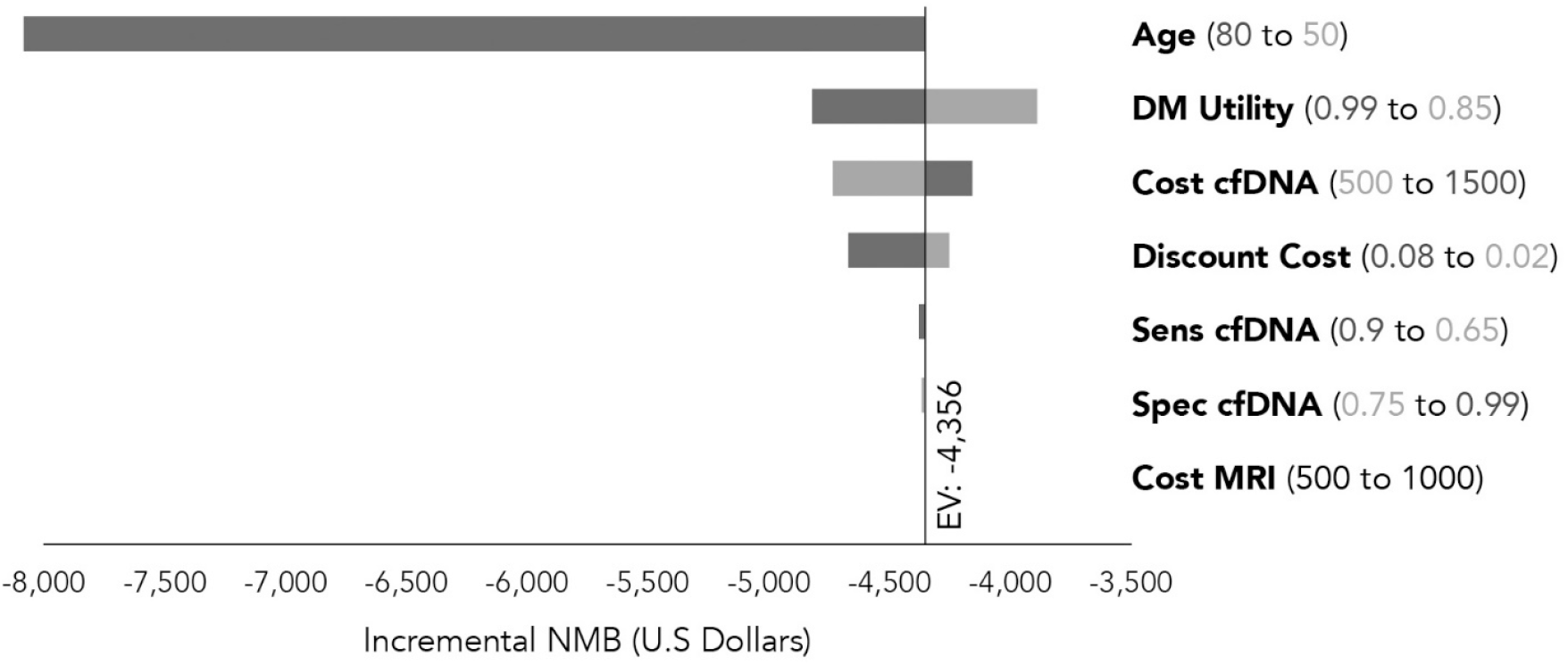
The Tornado diagram illustrates the results of one-way sensitivity analyses using important variables and the range of change in Incremental Net Medical Benefit (NMB) with each variable, with respect to the Willingness to Pay (WTP). Age at entry into the model has the most significant impact on NMB followed by health utility of diabetes and the cost of cfDNA test.

**Figure 3.**
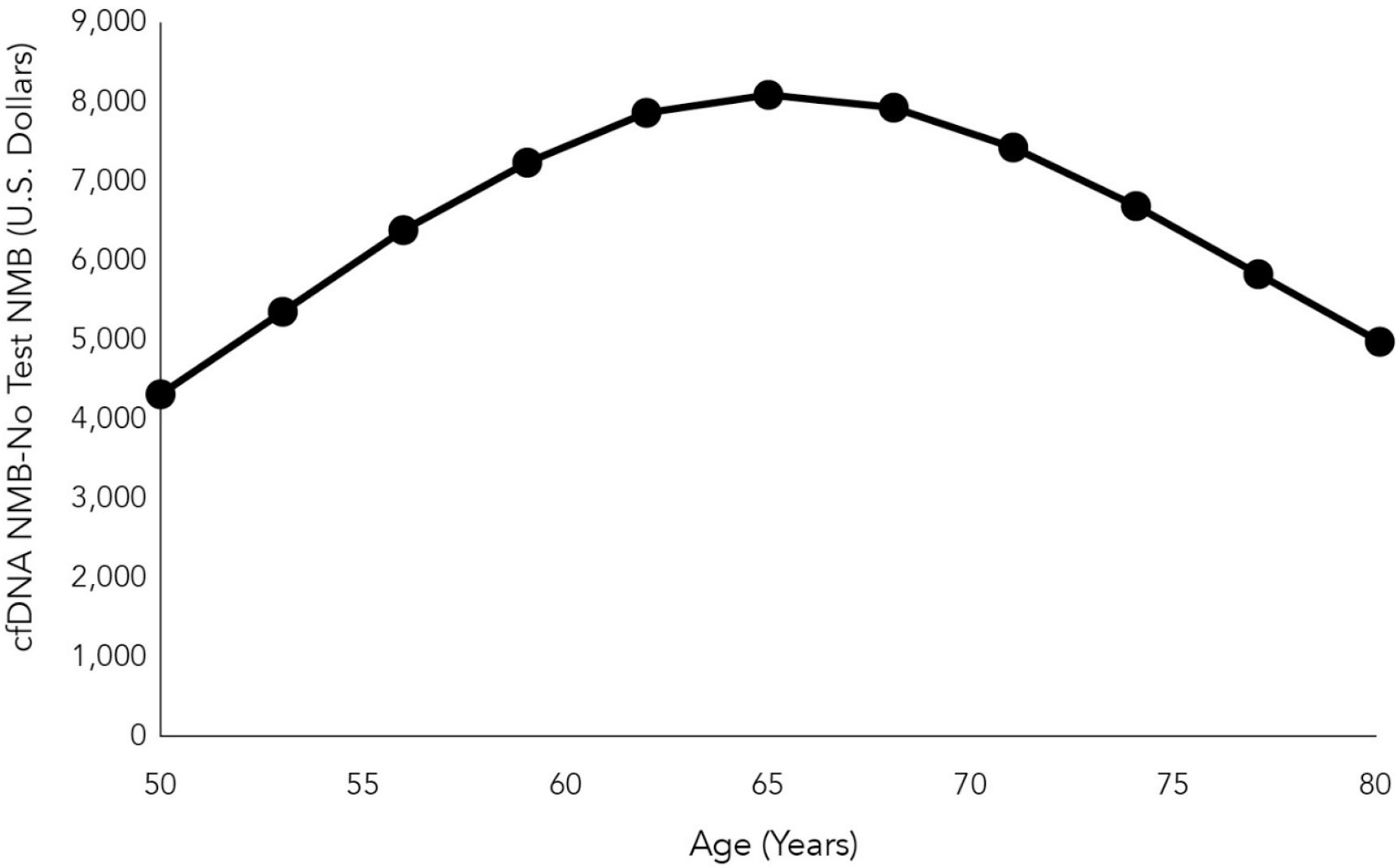
Result of a 1-way sensitivity analysis with the x-axis showing age at entry into the model and the y-axis showing the corresponding difference between Net Medical Benefit (NMB) in US dollars of the cfDNA test and the No Test strategy. The surveillance strategy with cfDNA testing consistently yields higher NMB than the No Test strategy, although for both strategies, overall NMB decreases with age at entry.

The two-way sensitivity analysis, which varies the cost of the cfDNA and MRI tests, further supports the preference for the cfDNA-based strategy in most scenarios, as illustrated in **Figure 4**.

**Figure 4.**
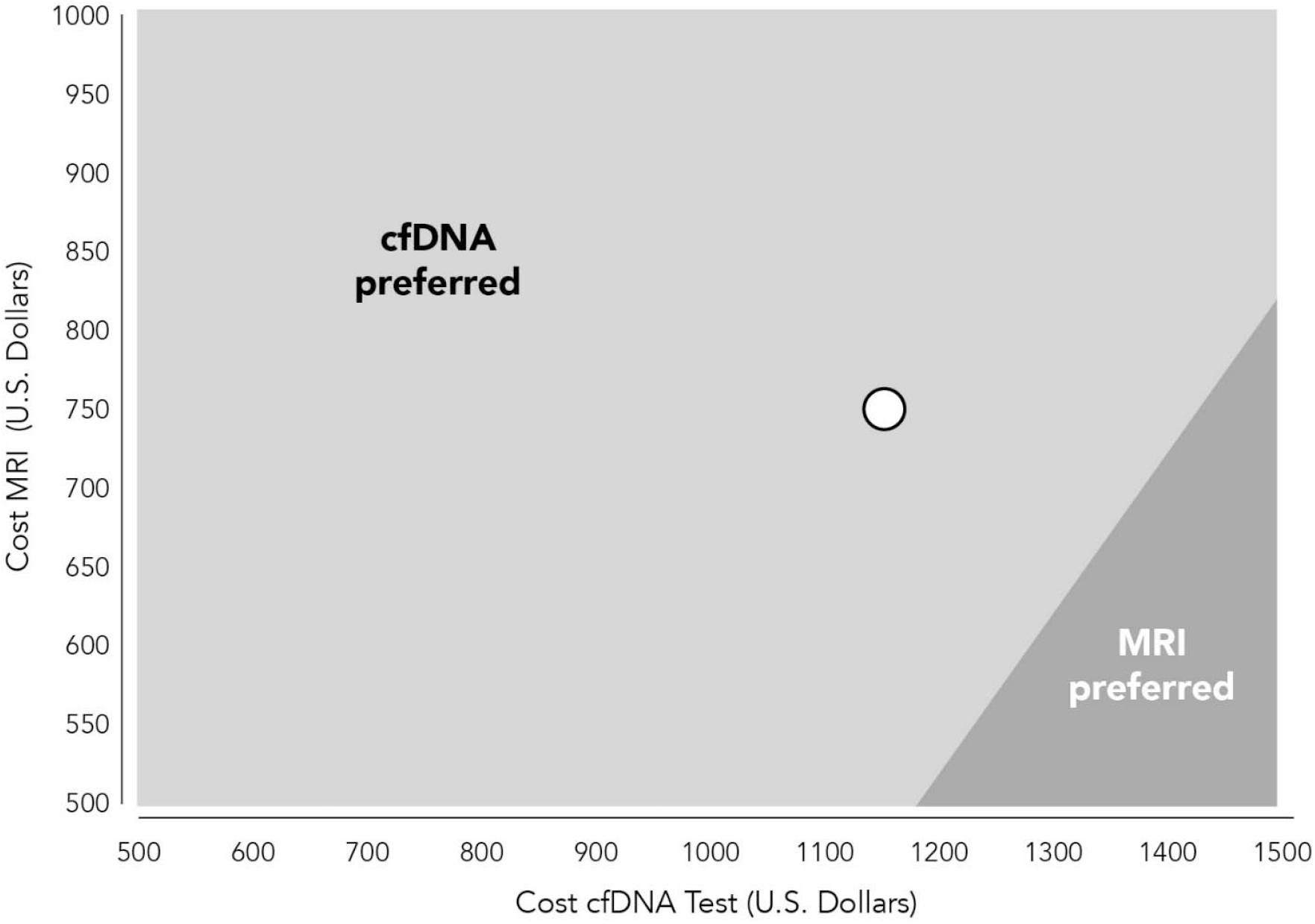
A 2-way sensitivity analysis with the cost of cfDNA testing along the X-axis and the cost of MRI along the Y-axis with the open circle representing the point estimates used in the baseline analysis. For a wide range of point estimates, including the baseline scenario, strategy of surveillance with cfDNA testing is preferred over MRI-based surveillance in terms of net benefit.

A second-order Monte Carlo simulation of a hypothetical cohort of 10,000 patients with new-onset diabetes revealed that the no-surveillance strategy led to 99 pancreatic cancer diagnoses (7.1% resectable) compared to 71 (32.4% resectable) in the cfDNA-tested group. Relative to no testing, cfDNA-based surveillance significantly reduced the risk of pancreatic cancer (Relative Risk, RR = 0.71; 95% CI, 0.53-0.97), and the number needed to test to prevent one case was 357 (95% CI, 187-3911). Similarly, for the diagnosis of pancreatic cancer at a resectable stage, with strategy II the estimated RR was 0.73 (95% CI, 0.61-0.86; p =0.002) and the number needed to test was 4 (95% CI, 2.8-6.9)

Strategies II and IV remained cost-effective across various willingness-to-pay values in most simulated scenarios. The acceptability curve from the Monte Carlo analysis (Figure 5) underscores this, showing that Strategy II is cost-effective in a high proportion of simulations based on willingness to pay.

**Figure 5.**
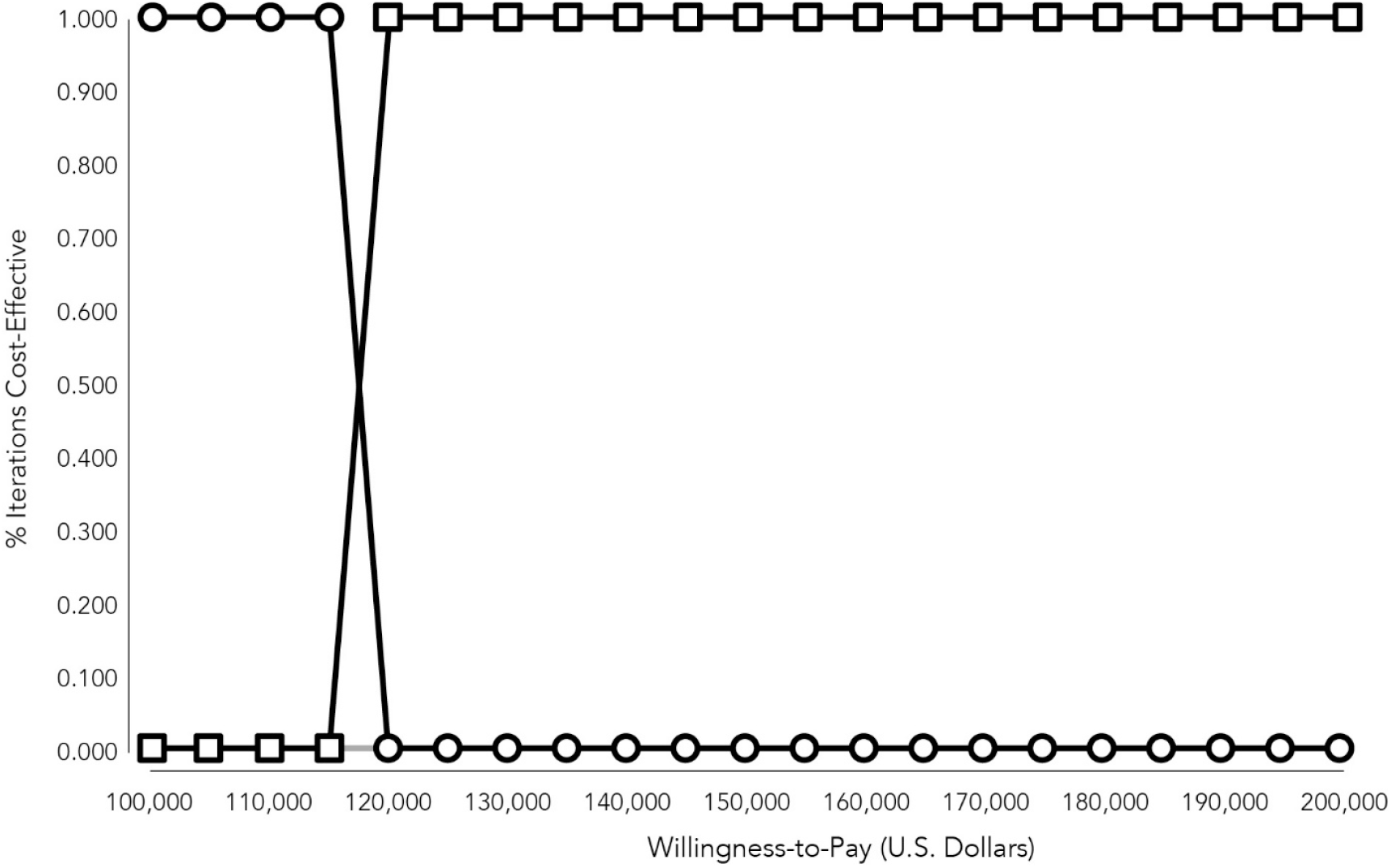
Cost-effectiveness Acceptability graph summarizes the results of the Monte Carlo simulations; above a WTP of approximately $115,000, the majority of the iterations favor the cfDNA-based strategy over the MRI-based strategy. Circles depict NOD surveillance using MRI, Squares depict NOD surveillance using the cfDNA test; the light gray line indicates the No Testing strategy.

## DISCUSSION

Risk stratification of NOD patients using the END-PAC PC risk algorithm, followed by testing with the cfDNA (Avantect) test, is an attractive strategy to reduce the burden of PC in this patient population. Previous analyses evaluated imaging modalities to test for PC in risk-stratified NOD patient populations. To our knowledge, the current study is the first to evaluate the economics of PC testing in the NOD population using a cfDNA-based test.

Currently, patients are assessed for PC using imaging-based tests, such as endoscopic ultrasound (EUS), abdominal CT scans, endoscopic retrograde cholangiopancreatography (ERCP), and magnetic resonance cholangiopancreatography (MRCP).^4^ Serum biomarkers, such as CA19-9 and the carcinoembryonic antigen (CEA), are used with imaging to evaluate symptomatic patients.^4^

However, no commonly used approach exists for early PC detection in NOD patients. Although effective, imaging modalities are not suitable for testing large patient populations; some communities may also have limited access to imaging. On the other hand, testing using cancer biomarkers such as CA19-9 and CEA is not recommended because of their lack of satisfactory sensitivity and specificity.^5^ In contrast, cfDNA analysis in peripheral blood is effective for early PC detection. The noninvasive nature of the cfDNA test also makes it attractive for use in large patient populations, especially those with limited access to imaging.

This study created a Markov model to compare several testing strategies, i.e., no-surveillance, surveillance using imaging modalities, and surveillance of NOD patients using a blood-based cell cfDNA epigenomic test. Using criteria from Sharma *et al*. (END-PAC), approximately 20% of the NOD patients were considered to be at higher risk for PC.^19^ The baseline analysis shows that the cfDNA-based testing strategy of a risk-stratified (END-PAC ≥3) NOD cohort is the most cost-effective compared to the standard of care (no testing for PC). CfDNA testing remains the preferred strategy when compared to imaging-based surveillance approaches. Although the MRI-based strategy was slightly lower in cost (CMS), it was a suboptimal strategy since it yielded lower QALY. This interpretation is further supported when the real-world out-of-pocket cost of the MRCP test ($3,600 to $6,160) is used.^35^

The findings presented here demonstrate that the cfDNA epigenomic test proves cost-effective in NOD patients at higher risk, with an ICER of $56,564 at a WTP of $100,000. Moreover, the model predicts that in a cohort of 10,000 patients with NOD, 1% would be diagnosed with PC in the no-surveillance strategy, with only 7.1% of cases treatable with surgery. In contrast, 71 PC cases are predicted to be detected with cfDNA testing. These cases would be more likely to be detected at an earlier, more treatable stage, with 32.4% predicted to be eligible for surgical resection.

Two previous studies have been published on the economics of testing NOD patients for PC.^20,21^ Schwartz *et al*. reported their findings using the END-PAC PC risk model to risk-stratify NOD patients. The authors developed an integrated decision tree and Markov state-transition model to evaluate the cost-effectiveness of PC testing in patients aged 50 years or older with NOD using CT imaging versus no testing.^21^ Their analysis predicted a cost per QALY gained of under $50,000 and under $100,000 in 34% and 99% of the simulations, respectively.^21^

Wang *et al*. compared PC early detection strategies targeting NOD patients aged ≥50 years at various minimal predicted PC risk thresholds (0.5% to 5.0%) vs standard of care in a Markov state–transition decision model. The authors used the risk stratification model published by Boursi and co-workers to define risk thresholds in the model population.^36^ In Wang *et al*.’s approach, the patients deemed to be at high risk underwent one-time diagnostic testing with abdominal MRI/magnetic resonance cholangiopancreatography (MRCP), and those with positive MRI underwent EUS/fine-needle aspiration (FNA). The low-risk group did not receive PC early detection testing (standard of care). The authors concluded that at a willingness to pay (WTP) threshold of $150,000 per quality-adjusted life-year, the early detection strategy targeting patients with a minimum predicted 3-year PC risk of 1% was cost-effective (ICER $116,911). At a WTP threshold of $100,000 per quality-adjusted life year, the early detection strategy at the 2% risk threshold was cost-effective (ICER: $63,045).

Both Schwartz et al. and Wang *et al*. agree that PC testing of NOD patients within three years of the diabetes diagnosis can be cost-effective, provided that the test is used in NOD patients at a higher risk for PC.

In contrast to the studies by Schwartz et al., which utilized contrast CT scans, and Wang et al., which employed MRCP as the primary surveillance modality in high-risk patients, our study primarily used the cfDNA blood test for surveillance. Despite differences in the surveillance tests, all three studies employed broadly comparable methodologies, model input estimates, and assumptions. Each study reached similar conclusions: a risk-stratified PC surveillance strategy is cost-effective in patients with NOD. Notably, the cfDNA blood test-based surveillance was found to be significantly more cost-effective, with an ICER of only $56,564, compared to $65,076 for contrast CT imaging (Schwartz *et al*.) and $116,911 per QALY for MRCP-based surveillance (Wang *et al*.).

## IMPLICATIONS AND FUTURE DIRECTIONS

According to the Centers for Disease Control and Prevention (CDC), over 1.2 million individuals receive a new diagnosis of T2D every year, with over 75% of them over the age of 45 years.^37^ Based on the END-PAC model, approximately 20% of these individuals would have an END-PAC score of 3 or higher, making them good candidates for cfDNA PC testing.^19^

The United States Preventive Services Task Force (USPSTF; 2019) recommends against surveillance for PC in the general population.^38^ The USPSTF explicitly recommends against testing NOD patients for PC.^38^ However, this recommendation is based primarily on the assumption that there is no sensitive and accurate test amenable to use in a large number of individuals. The availability of non-invasive, sensitive, and accurate testing modalities, such as the cfDNA (Avantect) test, is expected to have a significant impact during the next round of evidence evaluation by the USPSTF. This would be a welcome development, as evidence accumulated for the last 15 years shows that NOD patients are at a significantly higher risk of PC compared to the general population.^2^ This change in surveillance recommendations would be expected to have a substantial impact on the stage of detection and survival, as demonstrated by recently published data on the impact of active surveillance of individuals at high-risk for pancreatic cancer due to genetic predisposition and/or family history.^6^

Similar to other published economic analyses of pancreatic cancer surveillance in the NOD population, our study has certain inherent limitations. In the absence of published outcomes data on patients with NOD and PC detected by surveillance, we relied on data from surveillance programs targeting genetically predisposed high-risk individuals. Additionally, the performance characteristics of the novel cfDNA test have yet to be validated in a large cohort of patients with NOD.

Despite these limitations, our economic analysis also possesses several methodological strengths. To address the inherent uncertainty in the validity of input parameters, we conducted a comprehensive search of all published data. We used the most plausible range of estimates for sensitivity analyses. Furthermore, we employed advanced second-order Monte Carlo simulation techniques to account for these uncertainties, simulating real-life clinical practice and deriving robust statistical measures of effectiveness, such as relative risk and number needed to treat (NNT). We also applied the concepts of incremental net health benefit and willingness to pay, which are considered more appropriate indicators of cost-effectiveness than the ICER in analyses from a third-party payer perspective.

## CONCLUSIONS

This economic analysis indicates that cfDNA testing of risk-stratified NOD patients within three years of diabetes diagnosis is cost-effective. Clinical data are needed to confirm the impact of cfDNA testing on earlier PC cancer detection, mortality, and morbidity rates in the NOD population. However, the test’s analytical performance and predicted impact on early-stage pancreatic cancer detection, in conjunction with its cost-effectiveness, make it an attractive, non-invasive alternative testing modality for the surveillance of NOD patients.

## Data Availability

All data produced in the present study are available upon reasonable request to the authors

